# Mortality risk in infants receiving therapeutic care for malnutrition: A secondary analysis

**DOI:** 10.1101/2023.08.23.23294473

**Authors:** Imteaz Mahmud, Benjamin Guesdon, Marko Kerac, Carlos S. Grijalva-Eternod

## Abstract

**Background:** Small and nutritionally at-risk infants aged <6 months are at high risk of death, but important evidence gaps exist on how to best identify them. We aimed to determine associations between anthropometric deficits and mortality among infants <6m admitted to inpatient therapeutic care.

**Methods:** A secondary analysis of 2002-2008 data included 5,034 infants aged <6m from 12 countries. The prevalence, concurrence, and severity of wasted, stunted, underweight, and the Composite Index of Anthropometric Failure (CIAF) were analysed. We used logistic regression to examine the association of different anthropometric deficits with in-programme mortality.

**Results:** Among 3,692 infants aged <6m with complete data, 3,539 (95.8%) were underweight, 3,058 (82.8%) were wasted, 2,875 (77.8%) were stunted, and 3,575 (96.8%) had CIAF. Infants with multiple anthropometric deficits were more severely wasted, stunted, and underweight. A total of 141 infants died during inpatient therapeutic care. Among these, severely wasted (116) and severely underweight (138) infants had higher odds of mortality than normal infants (OR=2.1, 95% CI: 1.2-2.7, p=0.009, and OR=3.3, 95% CI: 0.8-13.6, p=0.09, respectively). Boys had higher odds of inpatient mortality than girls (OR=1.40, 95% CI: 1.02-1.92, p=0.03).

**Conclusion:** Multiple anthropometric deficits (CIAF) is common among infants <6m. Future work needs to explore which are the most useful indicator for programme admission and in-programme prognosis: our data supports both WLZ and WAZ, but future work which better accounts for admission bias is urgently needed. Boys appear to be most at-risk. Programmes should ensure that all infants receive timely, evidence-based, effective care.

## 1 INTRODUCTION

Child undernutrition, in all its manifestations, is a global health concern. According to the UNICEF/WHO/World Bank Group-joint child malnutrition estimates 2023 edition, around 148.1 million (22.3%) children under-five years of age are stunted (length-for-age z-score (LAZ) <-2), and 45 million (6.8%) are wasted (weight-for-length/height z-score (WLZ) <-2) (United Nations Children’s Fund (UNICEF), 2023). Annually, wasting and stunting are associated with around two million child deaths (Black et al., 2013). The long-term adverse effects of child undernutrition are also recognized: impaired neurocognitive development (Kirolos et al., 2022) and higher risks of non-communicable diseases in adult life (Grey et al., 2021). Among these undernourished children, infants under-six months of age (<6m) are especially vulnerable, with a global estimate of 20.1% infants <6m (23.8 million) underweight, 21.3% (24.5 million) wasted, and 17.6% (21.5 million) stunted (Kerac et al., 2021). Managing these small and nutritionally at-risk infants <6m is increasingly being recognised as an important challenge in child nutrition and health (Kerac & McGrath, 2017). Despite its significance, fundamental questions remain, notably on the optimal approach to identify at-risk infants <6m (Angood et al., 2015; Hoehn et al., 2021).

Anthropometry is an important, albeit imperfect, measure of nutritional status (Kerac et al., 2020). Current country-level child malnutrition guidelines are based on 2013 WHO guidelines which focus on WLZ <-2 (wasted) as the key anthropometric indicator to identify at-risk infants <6m (WHO, 2013). Limitations of this WLZ-based approach have been increasingly highlighted, including that: this index cannot be calculated in infants with lengths <45 cm; measuring length is time-intensive and logistically and technically difficult (Group & de Onis, 2006; Mwangome et al., 2012); the quality of length-based indicators is especially problematic in this age group, e.g., anthropometric data from infants <6m admitted to inpatient therapeutic centres was of lower quality than that of older children (Grijalva-Eternod et al., 2017). These challenges led to debates on whether other anthropometric indicators might be better alternatives, especially given that children under-five years of age with multiple anthropometric deficits, i.e., concurrently wasted, stunted, and underweight, were found to have the highest mortality risk (McDonald et al., 2013), with low weight-for-age z-score (WAZ) suggested as a better indicator (Khara et al., 2023; Myatt et al., 2018). A recent systematic review found that both WAZ and mid-upper arm circumference (MUAC) are better than WLZ at identifying infants <6m at the highest risk of mortality (Hoehn et al., 2021). Recognising these issues, just released July 2023 WHO guidelines now also recommend low WAZ and low MUAC alongside low WLZ as malnutrition programme admission criteria for infants <6m (WHO, 2023). However, the evidence underlying these recommendations is low quality: more data is needed, especially to inform country-level rollout. Related questions on which malnutrition programme admission criteria best identify the highest risk infants are around boys/girls differences: recent evidence suggests that boys, especially younger boys are more vulnerable than girls to being malnourished (Thurstans et al., 2022).

This study aimed to contribute evidence to decisions regarding the optimal identification of at-risk infants <6m by undertaking a secondary analysis of previously published data (Grijalva-Eternod et al., 2017). Our study objectives were threefold: (1) to describe the prevalence, concurrence, and severity of different anthropometric deficits in infants <6m admitted to therapeutic feeding programmes, (2) to explore the association between these deficits and mortality during inpatient therapeutic care, and (3) to explore sex differences in mortality among these infants <6m.

### Key messages

- Infants aged <6m with multiple anthropometric deficits present more severely wasted, stunted, and underweight. Prevalence of concurrent deficits increases with age within this group.
- Low WLZ and WAZ are most strongly associated with mortality risk in infants receiving inpatient therapeutic care. Associations between mortality and low LAZ are weakest, and LAZ also had a higher percentage of missing and flagged data.
- Boys had a higher prevalence of anthropometric deficits and a higher mortality risk than girls.
- Since our data are from infants <6m already admitted to therapeutic feeding programmes, future work is needed to see if these risk profiles also apply to the general population, or to infants <6m admitted to care using other criteria.

## 2 METHODS

### 2.1 Study design

We undertook a secondary analysis of fully anonymised routinely collected data from programmes providing malnutrition care to children aged under-five years. These programmes were conducted by registered organizations operating in countries with local permissions.

### 2.2 Data sources, settings, and participants

Details of the datasets used in this study are fully described elsewhere (Grijalva-Eternod et al., 2017). Briefly, 30 datasets from Action Contre la Faim were received following a data appeal. These datasets contained inpatient therapeutic data of individuals enrolled between 2002 and 2008 from 34 different field sites located in 12 countries, namely Afghanistan, Burundi, the Democratic Republic of the Congo, Ethiopia, Kenya, Liberia, Myanmar, Niger, Somalia, Sudan, Tajikistan, and Uganda (see **Table S1**). The criteria for children admitted to these centres were (1) infant too weak or feeble to suckle effectively, and/or (2) not gaining weight at home (by serial measurement of weight during growth monitoring), and/or (3) WLZ <-3, and/or (4) presenting bilateral pitting oedema. Data at admission and discharge were routinely collected at these field sites. The combined dataset contained data of 5,034 infants <6m that were used for this analysis.

### 2.3 Variables

The variables used for this study were age, sex, weight, and length at admission, and the discharge outcomes. Our primary outcome of interest was death as a discharge outcome. Length and weight were measured in cm and kg, respectively, with one decimal place precision.

We used Stata 16 software (StataCorp, 2019) for analysis. We calculated the WLZ, LAZ, and WAZ indices from age, sex, and anthropometric data, based on the 2006 WHO Child Growth Standards (WHO, 2006), using the *zscore06* Stata command (Leroy, 2011).

We flagged and excluded anthropometric indices as outliers using the 1995 WHO flexible cleaning criteria (Crowe et al., 2014) as follows: WLZ <-4 or >+4, LAZ <-4 or >+3, and WAZ <-4 or >+4 from the observed means. We removed from analysis all infants <6m with missing data on anthropometric indices (due to missing data or flagged values), sex, or discharge outcome.

We defined infants <6m as wasted, stunted, or underweight if their anthropometric indices WLZ, LAZ, and WAZ were <-2, respectively. We defined the Composite Index of Anthropometric Failure (CIAF) as all infants <6m that were either underweight, stunted, or wasted, and we generated the following six CIAF subcategories: (1) wasted only, (2) wasted and underweight, (3) wasted, stunted and underweight, (4) stunted and underweight, (5) stunted only, and (6) underweight only (see **Table S2**) (Nandy & Svedberg, 2012). There was no “wasted and stunted” category, as all children who meet these criteria were also underweight (Myatt et al., 2018).

### 2.4 Study size

We did not conduct *a priori* calculation of sample size: our sample was constrained by the number of children available in the compiled datasets.

### 2.5 Statistical analysis

We used descriptive statistics to assess the age and sex distributions of wasted, stunted, underweight, and CIAF prevalence. In each CIAF subcategory, we estimated the proportion of infants <6m discharged as dead. We estimated the mean anthropometric z-score values across different CIAF subcategories. We used the test of trend analysis to examine the linear age trends of anthropometric deficits. Finally, we used logistic regression analysis to estimate the mortality risk of different anthropometric deficits at admission and to explore its sex differences.

### 2.6 Ethical Statement

The Research Ethics Committee at the London School of Hygiene & Tropical Medicine Ethical approved this study (reference: 21985).

## 3 RESULTS

### 3.1 Study participants

Figure 1 shows the participant’s flow chart. A total of 885 infants <6m (17.5%) had any missing data and were excluded from the analysis. Of the anthropometric data, length was more commonly missing (5%) than weight (0.4%). Many infants <6m had missing anthropometric indices: 827 (16.4%) had missing WLZ, of which 564 (11.2%) had lengths <45 cm, 266 (5.3%) had missing LAZ, and 32 (0.6%) had missing WAZ. An additional 85 (1.7%) participants had missing discharge outcomes. On average, infants with no missing values were 1.5 months older than infants with missing values. Stratified by sex, 466 (19%) girls and 415 (16%) boys had any missing data. A total of 457 (9.1%) infants <6m had at least one anthropometric index flagged as an outlier, LAZ being the most flagged in 325 infants (7.8%). After excluding those with missing or flagged data, 3,692 (74.5%) infants <6m were included for analysis.

**FIGURE 1:**
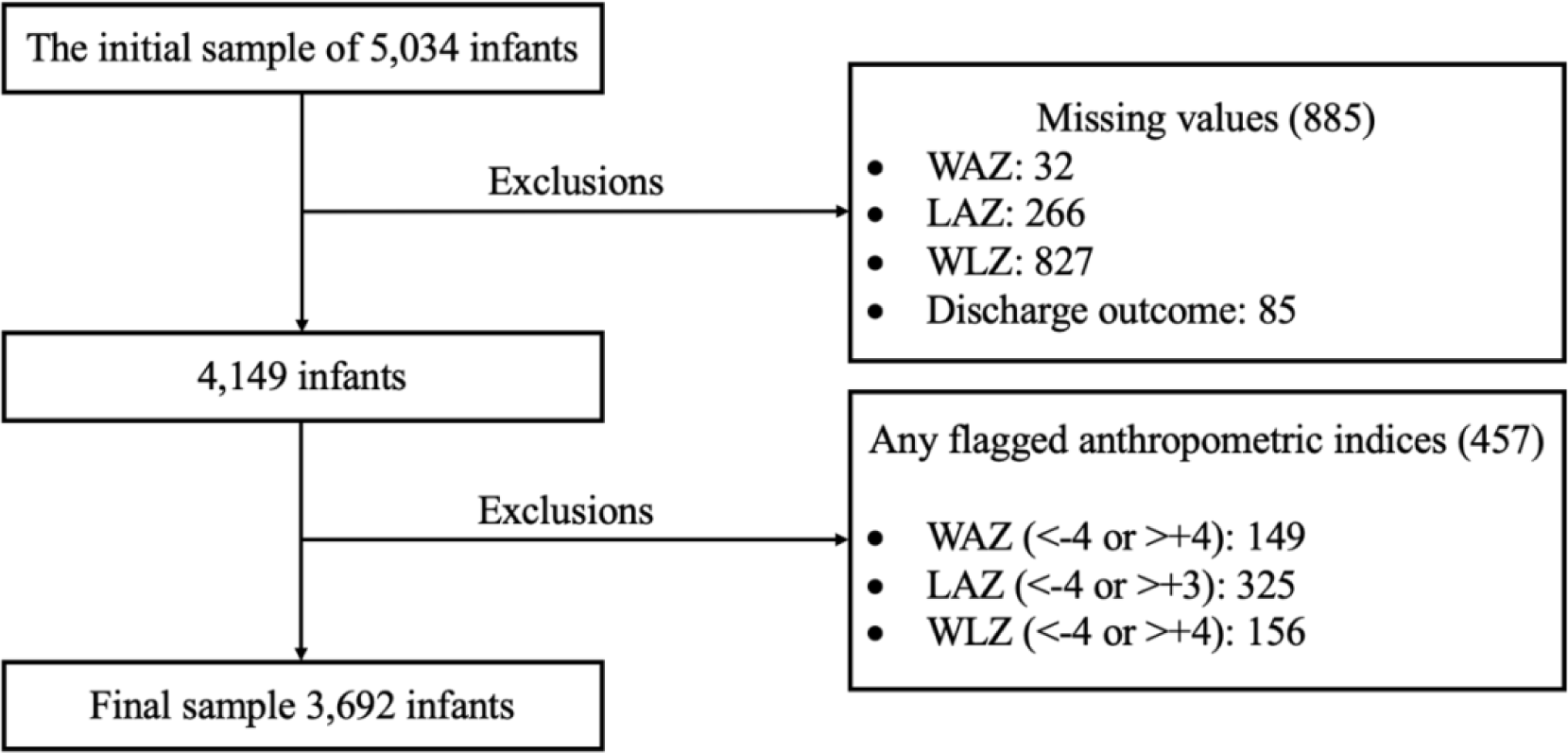
Study participant’s flow chart

### 3.1 Age and sex distribution of anthropometric deficits at admission

**Table 1** shows the age distribution of different anthropometric deficits, CIAF, and subcategories at admission. Underweight was the most prevalent anthropometric deficit affecting 3,539 infants (95.9%), 3,058 (82.8%) and 2,875 (77.9%) infants were wasted and stunted, respectively. The mean age of wasted, stunted, and underweight infants were comparable.

**TABLE 1.**
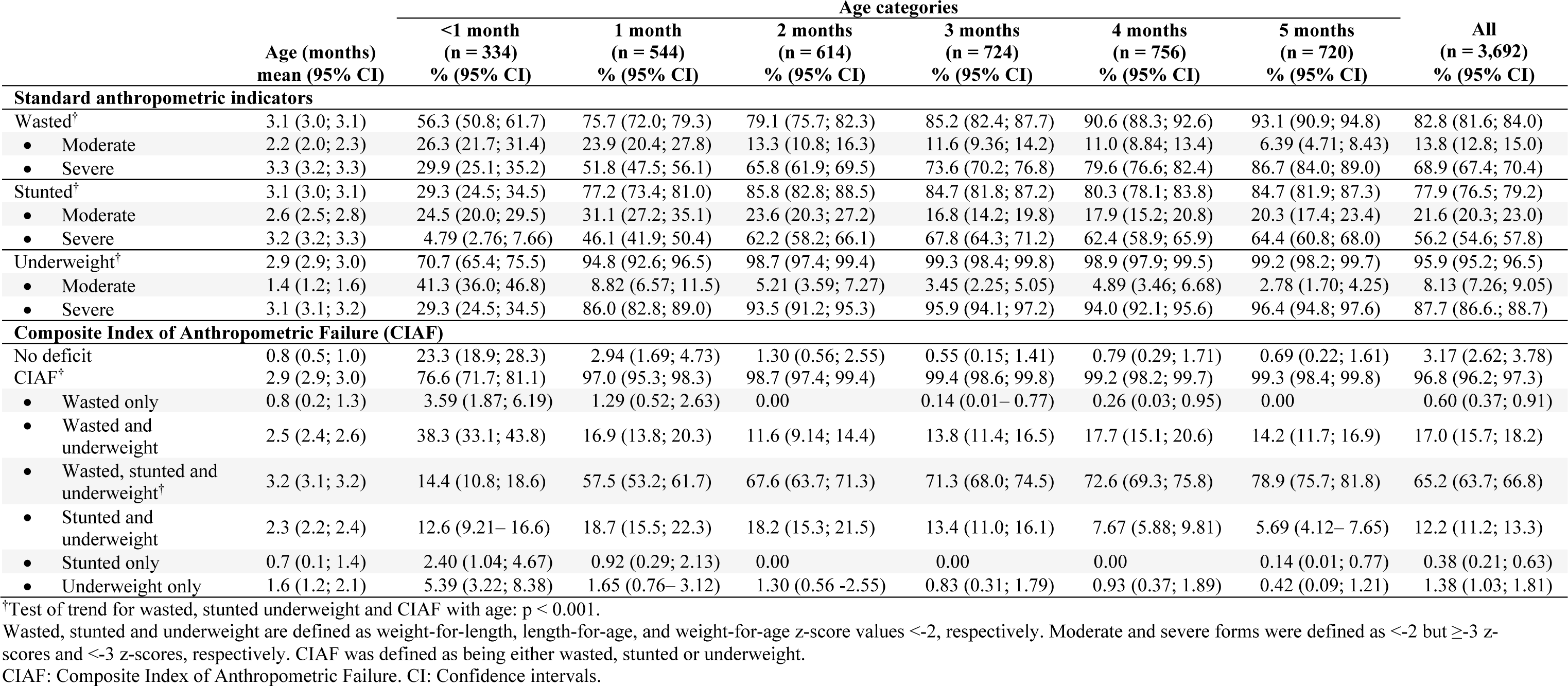
Descriptive statistics of different anthropometric deficits.

The CIAF indicator captured 3,575 (96.8%) of the admitted infants <6m. Only 3.2% of infants <6m had no anthropometric deficit at admission, and only 2.3% presented with single anthropometric deficits, namely wasted only, stunted only, or underweight only. Most infants <6m (65.2%) presented with all three concurrent anthropometric deficits (wasted, stunted, and underweight) (see Figure 2). Infants with multiple anthropometric deficits were older than infants with single deficits. The prevalence of different anthropometric deficits increased with age, whereby infants aged <1 month had the lowest CIAF prevalence. The test of trend showed a linear trend with age for both wasted, stunted, underweight, and CIAF prevalence (p<0.001). Boys had a higher prevalence of multiple anthropometric deficits compared to girls, with 68.2% (95% CI: 66.0-70.3) of wasted, stunted, and underweight boys compared to 62.0% (95% CI: 59.7-64.3) of girls (see **Table S3**).

**FIGURE 2:**
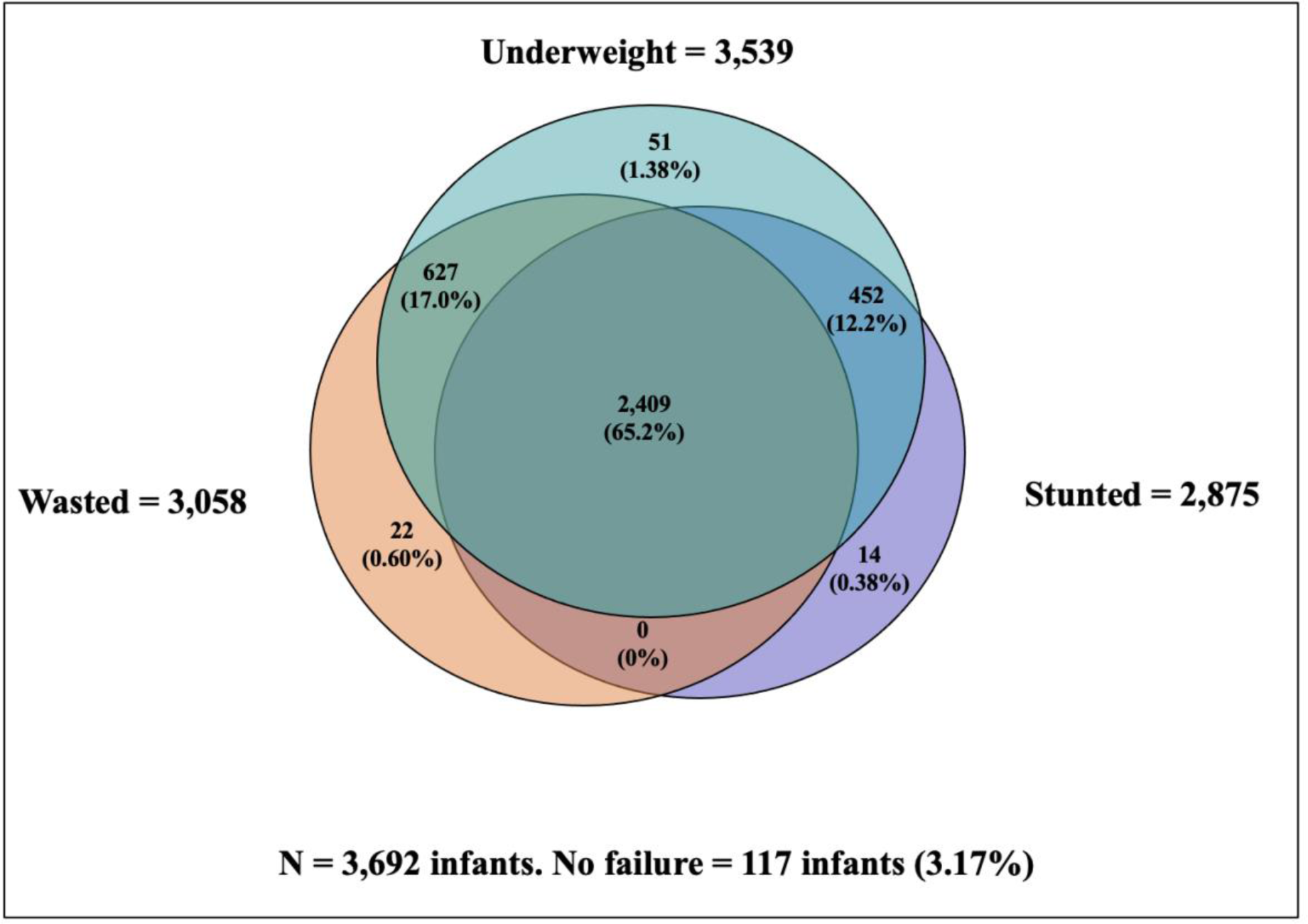
Venn diagram showing the prevalence and overlap of different anthropometric deficits.

### 3.1 The severity of anthropometric deficits

**Table 2** shows the mean values of WAZ, WLZ, and LAZ in the different CIAF subcategories. Infants <6m with multiple anthropometric deficits had the lowest mean values of the anthropometric indices. Infants who were concurrently wasted, stunted, and underweight had –5.51, –4.20, and –3.91 mean values of WAZ, WLZ, and LAZ, respectively.

**TABLE 2.** Mean anthropometric z-score values by Composite Index of Anthropometric Failure (CIAF) category (n = 3,692).

|  | WAZ<br>Mean (95% CI) | WLZ<br>Mean (95% CI) | LAZ<br>Mean (95% CI) |
| --- | --- | --- | --- |
| No deficit (n = 117) | -1.42 (-1.50; -1.34) | -0.91 (-1.03; -0.80) | -1.03 (-1.12; -0.93) |
| CIAF (n = 3,575) | -4.93 (-4.97; -4.89) | -3.75 (-3.80; -3.70) | -3.40 (-3.45; -3.35) |
| • Wasted only (n = 22) | -1.89 (-1.96; -1.82) | -2.42 (-2.51; -2.32) | -0.50 (-0.58; -0.41) |
| • Wasted and underweight (n = 627) | -3.71 (-3.78; -3.64) | -4.27 (-4.37; -4.17) | -1.26 (-1.30; -1.22) |
| • Wasted, stunted and underweight (n = 2,409) | -5.51 (-5.55; -5.47) | -4.20 (-4.24; -4.15) | -3.91 (-3.96; -3.87) |
| • Stunted and underweight (n = 452) | -4.05 (-4.14; -3.95) | -1.10 (-1.16; -1.04) | -4.03 (-4.14; -3.91) |
| • Stunted only (n = 14) | -1.89 (-1.94; -1.85) | -0.07 (-0.23; 0.10) | -2.30 (-2.41; -2.19) |
| • Underweight only (n = 51) | -2.33 (-2.41; -2.26) | -1.56 (-1.64; -1.49) | -1.64 (-1.70; -1.57) |

Wasted, stunted, and underweight are defined as weight-for-length, length-for-age, and weight-for-age z-score values <-2, respectively. CIAF was defined as either wasted, stunted or underweight.
CIAF: Composite Index of Anthropometric Failure. WAZ: Weight-for-age z-score. WLZ: Weight-for-length z-score. LAZ: Length-for-age z-score. CI: Confidence intervals.

### 3.1 Mortality risk associated with anthropometric deficits

**Table 3** shows the proportion of infants <6m who died whilst receiving programme care, organised by different anthropometric deficits. A total of 141 (3.8%) infants <6m died while receiving inpatient therapeutic care. Of the commonly used stand-alone anthropometric indicators, severely underweight captured the highest number of infants <6m (n=138) who died, followed by severely wasted (n=116) and then severely stunted (n=88).

**TABLE 3:** Association of different anthropometric deficits with mortality during inpatient therapeutic care, infants under 6 months (N = 3,692, total deaths = 141)

|  | N (%) | Mortality |  |  |
| --- | --- | --- | --- | --- |
|  |  | Deaths (%) | Odds ratio (95% CI) | p-value |
| Standard anthropometric indicators |  |  |  |  |
| Not wasted | 634 (17.1) | 14 (2.2) | 1.0 (ref.) |  |
| Wasted | 3,058 (82.8) | 127 (4.1) | 1.9 (1.1 – 3.3) | 0.02 |
| • Moderate | 513 (13.8) | 11 (2.1) | 0.9 (0.4 – 2.1) | 0.94 |
| • Severe | 2,545 (68.9) | 116 (4.5) | 2.1 (1.2 – 3.7) | 0.009 |
| Not Stunted | 817 (22.1) | 31 (3.7) | 1.0 (ref.) |  |
| Stunted | 2,875 (77.8) | 110 (3.8) | 1.0 (0.6 – 1.5) | 0.96 |
| • Moderate | 799 (21.6) | 22 (2.7) | 0.7 (0.4 – 1.2) | 0.24 |
| • Severe | 2,076 (56.2) | 88 (4.2) | 1.1(0.7 – 1.7) | 0.58 |
| Not Underweight | 153 (4.1) | 2 (1.3) | 1.0 (ref.) |  |
| Underweight | 3,539 (95.8) | 139 (3.9) | 3.0 (0.7 – 12.5) | 0.11 |
| • Moderate | 300 (8.1) | 1 (0.3) | 0.2 (0.02 – 2.8) | 0.26 |
| • Severe | 3,239 (87.7) | 138 (4.2) | 3.4 (0.8 – 13.6) | 0.09 |
| Composite index of anthropometric failure (CIAF) |  |  |  |  |
| No CIAF | 117 (3.1) | 2 (1.7) | 1.0 (ref.) |  |
| CIAF | 3,575 (96.8) | 139 (3.8) | 2.3 (0.5 – 9.5) | 0.24 |
| • No failure | 117 (3.1) | 2 (1.7) | 1.0 (ref.) |  |
| • Wasted only | 22 (0.6) | 0 |  |  |
| • Wasted and underweight | 627 (16.9) | 29 (4.6) | 2.8 (0.6 – 11.8) | 0.16 |
| • Wasted, stunted and underweight† | 2,409 (65.2) | 98 (4.0) | 2.4 (0.5 – 10.0) | 0.21 |
| • Stunted and underweight | 452 (12.2) | 12 (2.6) | 1.5 (0.3 – 7.10) | 0.56 |
| • Stunted only | 14 (0.3) | 0 |  |  |
| • Underweight only | 51 (1.3) | 0 |  |  |

Wasted, stunted, and underweight are defined as weight-for-length, length-for-age, and weight-for-age z-score values <-2, respectively. Moderate and severe forms were defined as <-2 but ≥-3 z-scores and <-3 z-scores, respectively. CIAF was defined as either wasted, stunted or underweight.
CIAF: Composite Index of Anthropometric Failure. CI: Confidence intervals

None of the infants <6m presenting single anthropometric deficits (wasted only, stunted only, or underweight only) at admission were dead at discharge. The proportion of deaths was greater among those with multiple anthropometric deficits.

The odds of dying during inpatient therapeutic care were significantly higher in severely wasted infants (OR=2.11, 95% CI: 1.20-3.70, p-value=0.009) compared to non-wasted infants. Infants with severe underweight had higher odds of inpatient mortality compared to non-underweight infants, albeit evidence for this was weak, statistically non-significantly (OR=3.35, 95% CI: 0.82-13.6, p-value=0.09). Severely stunted infants had non-significantly higher odds of dying than those non-stunted (OR=1.12, 95% CI=0.73-1.70; p-value=0.58).

Among the infants <6m that died, 85 (60.3%) were boys, and 56 (39.7%) were girls. Logistic regression showed that boys had significantly greater odds of mortality (OR=1.40, 95% CI: 1.02-1.92, p-value=0.03) (see **Table S4**).

## 4 DISCUSSION

Our main findings were that infants <6m who presented with multiple anthropometric deficits had the worst form of anthropometric profile and the highest mortality, albeit the latter was not statistically significant, compared to those with single or no anthropometric deficits. Wasted at admission was associated with dying during inpatient care. There was weak evidence that the odds of dying were also greater among those underweight. Finally, boys had a higher risk of mortality compared to girls.

In this analysis, we observed no inpatient deaths among infants <6m with single anthropometric deficits (wasted only, stunted only, or underweight only). Deaths clustered among infants with multiple forms of anthropometric deficit. This observation is in line with other studies in older children that found greater mortality risk among those who presented with more than one form of anthropometric deficit (McDonald et al., 2013; Myatt et al., 2018; Olofin et al., 2013). A simple explanation is that these infants had more severe anthropometric deficits compared with those having single or no deficits: this association between the severity of malnutrition and risk of death has long been recognised (Pelletier, 1994). Data in older children shows that malnutrition impacts negatively on muscle and fat mass (Briend et al., 1989). Infants with multiple anthropometric deficits might have lower muscle mass (Reeds et al., 1994) because of both malnutrition and shorter limbs. Whenever these infants <6m present an infection, their bodies might fail to supply the required amino acids to sustain the immune response and vital organ functions, which in turn increases the probability of death (Briend et al., 1989; Broeck et al., 1998). In addition, infants with recurrent infections may have reduced appetites and increased metabolic demands (Tomkins & Watson, 1989; WHO, 2015). This, in turn, might lead to poor breastfeeding and further growth faltering in a continuous vicious cycle. If these infants live in a context of poor sanitation, deprivation, and a lack of available healthcare services, they may fail to recover and ultimately die (Scrimshaw et al., 1968). This might also explain the higher vulnerability to death of infants with multiple anthropometric deficits (McDonald et al., 2013).

Given their heightened mortality risk and the severity of anthropometric profiles, infants with multiple anthropometric deficits might need to be prioritised to receive care promptly, as this might help reduce their mortality. However, implementing this might not be without challenges. CIAF and its subcategories introduce a new dimension of anthropometric deficits affecting infants, which might remain unnoticed when using conventional siloed nutrition indicators. CIAF could be understood as a single indicator presenting a more comprehensive picture of undernutrition and mortality risk at admission. Benefits might be obtained if CIAF and its subcategories are estimated and reported in malnutrition care programmes and nutrition surveys to better represent the anthropometric profile of the undernourished infant <6m population and to help further assess the association between anthropometric deficits and their associated risk among infants. Balancing potential benefits of routine CIAF assessment and reporting is its time demands and the added complexity to programmes without a clear guarantee of improved outcomes. Future research is needed to explore the added value of CIAF in other programmes/datasets and to understand if simplified approaches might be just as effective at identifying the highest-risk infants <6m (e.g., using WAZ or MUAC alone as suggested by a recent systematic review (Hoehn et al., 2021).

In our data, we observed that at admission, boys had a greater proportion who were concurrently wasted, stunted and underweight compared to girls and presented with a higher mortality risk. Two recent meta-analyses provide support to the nutritional disadvantages in boys, one conducted in 84 countries among older children aged 6-59 months (Khara et al., 2018), and another conducted in 74 studies among children aged 0-59 months (Thurstans et al., 2020). However, contrary to our results, the risk of death was found to be similar in boys and girls (Thurstans et al., 2023). The 2020 Global Nutrition Report also showed that boys under-five years of age had a higher prevalence of stunted and wasted compared to girls (Micha et al., 2020). Our study added to evidence that this difference in burden and associated mortality risk might be present at an earlier age. Many possible mechanisms might explain these sex differences (Thurstans et al., 2022). Possibly in response to greater biological vulnerability, boys are often prioritized in terms of parental treatment-seeking behaviours (Mahmud et al., 2020). Programmes might need to account for biological and social risk differentials to ensure that all children receive the care needed to survive and thrive.

In our dataset, we lacked full information on the reasons for admission although, at the time covered by the datasets, being wasted was the main anthropometric criteria for admission to programme care. Nonetheless, of those admitted, only 83% were wasted, 78% were stunted, whilst most were found to be underweight or CIAF (96% and 97%, respectively). Whilst it is likely that some infants were admitted for other reasons, like maternal concerns or feeding problems, our findings are supportive of calls for continuing exploration of using low WAZ to identify infants <6m at high-risk. Additional arguments found in the literature are (1) that low WAZ is most closely associated with risk of mortality (Hoehn et al., 2021), (2) weight is measured routinely in growth monitoring as well as therapeutic care programmes, so it does not add further burden to healthcare workers (de Onis et al., 2004), (3) WAZ can be used to assess infants <6m at-risk whose length is <45 cm, and (5) it is easier and quicker to assess than WLZ because it does not require length measurement.

We urge caution in interpreting our results. First, the lower proportion of wasted infants might reflect wider clinical criteria at admission, such as the inability to feed effectively, irrespective of anthropometric status. Second, because we only studied infants already admitted to therapeutic feeding, we cannot extrapolate our results to a wider population of infants <6m or make evidence-based statements about which criteria might be best for admitting to care. Further research on these issues is certainly needed using population-based samples or datasets. Complementary research like ours is also important to understand which infants – of those admitted – are the highest risk and might thus need different treatment or follow-up.

### 4.1 Future research

Our study was conducted among infants receiving inpatient therapeutic care. Future research should focus on the general population of infants <6m regarding the prevalence of anthropometric deficits, their concurrence, and their association with death. We found that boys had a higher prevalence of anthropometric deficits and their concurrence, and many possible mechanisms described elsewhere may explain this higher prevalence (Thurstans et al., 2022). Nonetheless, explanations for our findings remain unknown. Further research is needed to understand sex differences, and its implications for nutrition programmes in different settings. Though our data did not show the same statistically significant advantages of low WAZ over other indicators as others have found, more data from other settings on the question of “best” anthropometric indicators is needed. MUAC is another indicator that we did not have in our dataset that needs future investigation. Appropriate treatment protocols, follow-up, and discharge criteria also need to be researched. In this analysis, infants with multiple anthropometric deficits presented with more severe forms of malnutrition and appear to have greater, non-significant odds of dying. Longitudinal studies are needed to expand our understanding of the concurrent wasted and stunted processes in infants <6m.

### 4.2 Strengths and limitations

We acknowledge a number of limitations. We might have measurement-, seasonality-(Maleta et al., 2003), and selection bias affecting our findings, as the data was collected at different years and seasons, by various staff, in 34 different therapeutic centres from 12 countries, and merged for analysis. Our infants <6m sample were admitted to therapeutic care based on their vulnerability. Consequently, the patterns we observed in these infants might not be comparable to a broader infant <6m population. Our study might lack the power to confidently ascertain our findings as we undertook a secondary analysis. Furthermore, in some CIAF subcategories, the samples were small, and might have resulted in large confidence intervals in some estimates. We lacked information regarding co-morbidities (e.g., diarrhoea, HIV), congenital anomalies, maternal stature, weight gain during pregnancy, pre-natal or birth history (low birthweight or small for gestational age), or breastfeeding status of the participants, which might affect the observed patterns between anthropometric deficits and mortality (Black et al., 2013; De Onis et al., 2013; Rayco-Solon et al., 2005). Younger infants and girls had a higher rate of missing values, which might have incorporated age- and sex-bias. Infants <6m discharged or cured from the inpatient therapeutic care programme were not followed up, so the possibility of a recurrence or even death is unknown.

Balancing these limitations are study strengths. To our knowledge, this is the largest multi-centre analysis of inpatient therapeutic care data of infants <6m. As such, this dataset allows for a more global understanding of the management of acute malnutrition in this age group; and our findings might be generalisable to other therapeutic care settings. Given the paucity of data, this analysis adds to the evidence base. The WHO agenda is to revise the protocol to treat infants <6m. It will require consistent evidence across different settings, programmes, or countries.

## 5 CONCLUSION

A high proportion of infants <6m admitted to inpatient therapeutic care have multiple anthropometric deficits and appear to have worse nutritional status. CIAF and its subcategories might be useful to identify infants <6m at higher risk of dying. Of the stand-alone nutrition indicators, WLZ and WAZ at admission perform better than LAZ at identifying at-risk infants <6m. Further research is needed to understand the practical implications of using different indicators to identify and provide malnutrition care to infants <6m. In addition, research is needed to better understand the causes behind and possible responses to the nutrition disadvantage and higher mortality risk observed in boys at this early age.

## Data Availability

All data produced in the present study are available upon reasonable request to the authors

## Acknowledgements

We are thankful to Action Contre la Faim - France for providing the data used in this analysis.

## Contributions

IM, MK, and CSG-E conceived the study. BG, MK, and CSG-E contributed data. IM and CSG-E analysed and interpreted the data. IM wrote the initial draft of the manuscript. All authors contributed to the manuscript revisions. All authors read and approved the final manuscript.

