## Supplemental file for "Mortality risk in infants receiving therapeutic care for malnutrition: A secondary analysis"

**Table S1: Programme datasets by country**

| Country | Years | N (%) | Age (months) mean (95% CI) | Male sex N (%) |
|---|---|---|---|---|
| **Afghanistan** | 2002 – 2004 | 1,032 (20.5) | 3.1 (3.0 – 3.2) | 564 (54.7) |
| **Burundi** | 2006 – 2007 | 146 (2.90) | 2.4 (2.2 – 2.6) | 67 (45.8) |
| **DRC** | 2005 – 2007 | 2,240 (44.5) | 1.9 (2.6 – 3.5) | 1,106 (49.3) |
| **Ethiopia** | 2008 | 34 (0.68) | 3.1 (3.0 – 4.1) | 15 (45.4) |
| **Kenya** | 2005 – 2007 | 43 (0.85) | 3.2 (2.8 – 3.6) | 23 (53.4) |
| **Liberia** | 2006 – 2008 | 186 (3.69) | 2.7 (2.4 – 2.9) | 97 (52.1) |
| **Myanmar** | 2006 – 2008 | 182 (3.62) | 1.9 (1.7 – 2.1) | 83 (45.8) |
| **Niger** | 2006 – 2008 | 151 (3.00) | 2.3 (2.1 – 2.5) | 50 (51.0) |
| **Somalia** | 2006 – 2008 | 402 (7.99) | 3.9 (3.8 – 4.0) | 211 (52.4) |
| **Sudan** | 2006 – 2008 | 378 (7.51) | 2.0 (1.8 – 2.1) | 206 (54.5) |
| **Tajikistan** | 2005 – 2006 | 107 (2.13) | 3.6 (3.3 – 3.8) | 58 (54.2) |
| **Uganda** | 2005 – 2007 | 133 (2.64) | 3.6 (3.3 – 3.8) | 71 (53.3) |
| **Total** | 2002 – 2008 | 5,034 (100) | 2.5 (2.4 – 2.5) | 2,580 (51.2) |

**Table S2: Subgroups of the Composite index of anthropometric failure (CIAF)**

| Description | Wasted | Stunted | Underweight |
|---|---|---|---|
| **No deficits** | No | No | No |
| **Wasted only** | Yes | No | No |
| **Wasted and underweight** | Yes | No | Yes |
| **Wasted, stunted and underweight** | Yes | Yes | Yes |
| **Stunted and underweight** | No | Yes | Yes |
| **Stunted only** | No | Yes | No |
| **Underweight only** | No | No | Yes |

Wasted, stunted, and underweight are defined as weight-for-length, length-for-age, and weight-for-age z-score values <-2, respectively.

**Table S3: Sex distribution of different anthropometric failures**

| | Boys % | Girls % | All % | Sex ratio boys: girls |
|---|---|---|---|---|
| **Standard anthropometric indicators** | | | | |
| Wasted | 84.4 | 81.1 | 82.8 | 1: 0.96 |
| • Moderate | 12.5 | 15.4 | 13.8 | 1: 1.13 |
| • Severe | 71.9 | 65.7 | 68.9 | 1: 0.83 |
| Stunted | 80.4 | 75.1 | 77.8 | 1: 0.93 |
| • Moderate | 19.9 | 23.5 | 21.6 | 1: 1.08 |
| • Severe | 60.5 | 51.6 | 56.2 | 1: 0.78 |
| Underweight | 97.1 | 94.5 | 95.8 | 1: 0.97 |
| • Moderate | 7.62 | 8.68 | 8.13 | 1: 1.04 |
| • Severe | 89.5 | 85.8 | 87.7 | 1: 0.87 |
| **Composite index of anthropometric failure (CIAF)** | | | | |
| No deficits | 2.12 | 4.31 | 3.17 | 1: 1.85 |
| CIAF | 97.8 | 95.7 | 96.8 | 1: 0.98 |
| • Wasted only | 0.36 | 0.85 | 0.60 | 1: 2.14 |
| • Wasted and underweight | 15.8 | 18.2 | 16.9 | 1: 1.05 |
| • Wasted, stunted and underweight | 68.2 | 62.0 | 65.2 | 1: 0.83 |
| • Stunted and underweight | 11.8 | 12.8 | 12.2 | 1: 0.99 |
| • Stunted only | 0.41 | 0.34 | 0.38 | 1: 0.75 |
| • Underweight only | 1.30 | 1.48 | 1.38 | 1: 1.04 |

Wasted, stunted, and underweight are defined as weight-for-length, length-for-age, and weight-for-age z-score values <-2, respectively. Moderate and severe forms were defined as <-2 but ≥-3 z-scores and <-3 z-scores, respectively. CIAF was defined as either wasted, stunted or underweight.

**Table S4: Sex difference in mortality**

| | | Mortality | | | |
|---|---|---|---|---|---|
| **Sex** | **N** | **Deaths** | **% (95% CI)** | **Odds ratio (95% CI)** | **P-value** |
| **Female** | 1,762 | 56 | 3.18 (2.41; 4.11) | Ref. | |
| **Male** | 1,930 | 85 | 4.40 (3.53; 5.42) | 1.40 (1.02; 1.92) | 0.03 |
